# Clinical activity of Mitogen-Activated Protein Kinase (MAPK) inhibitors in patients with MAP2K1 (MEK1)-mutated metastatic cancers

**DOI:** 10.1101/2024.03.23.24304779

**Authors:** Matthew Dankner, Emmanuelle Rousselle, Sarah Petrecca, François Fabi, Alexander Nowakowski, Anna-Maria Lazaratos, Charles Vincent Rajadurai, Andrew J. B. Stein, David Bian, Peter Tai, Alicia Belaiche, Meredith Li, Andrea Quaiattini, Nicola Normanno, Maria Arcila, Arielle Elkrief, Douglas B. Johnson, Marc Ladanyi, April A. N. Rose

## Abstract

**PURPOSE:** MAP2K1/MEK1 mutations are potentially actionable drivers in cancer. MAP2K1 mutations have been functionally classified into three groups according to their dependency on upstream RAS/RAF signaling. However, the clinical efficacy of MAPK pathway inhibitors (MAPKi) for MAP2K1 mutant tumors is not well defined. We sought to characterize the genomic and clinical landscape of MAP2K1 mutant tumors to evaluate the relationship between MAP2K1 mutation Class and clinical activity of MAPKi.

**METHODS:** We interrogated AACR GENIE (v13) to analyze solid tumors with MAP2K1 mutations. We performed a systematic review and meta-analysis of published reports of patients with MAP2K1 mutant cancers treated with MAPKi according to PRISMA guidelines. The primary endpoint was progression-free survival (PFS), and secondary endpoints were overall response rate (ORR), duration of response (DOR), and overall survival (OS).

**RESULTS:** In the AACR GENIE dataset, Class 2 MAP2K1 mutations (63%) were more prevalent than Class 1 (24%) and Class 3 (13%) mutations (P<0.0001). Co-occurring MAPK pathway activating mutations were more likely to occur in Class 1 versus Class 2 or 3 MAP2K1 mutant tumors (P<0.0001). Our systematic meta-analysis of the literature identified 46 patients with MAP2K1 mutant tumors who received MAPKi. In these patients, ORR was 28% and median PFS was 3.9 months. ORR did not differ according to MAP2K1 mutation class or cancer type. However, patients with Class 2 mutations experienced longer PFS (5.0 months) and DOR (23.8 months) compared to patients with Class 1, 3 or unclassified MAP2K1 mutations (PFS 3.5 months, P=0.04; DOR 4.2 months, P=0.02).

**CONCLUSION:** Patients with Class 2 MAP2K1 mutations represent a novel subgroup that may derive benefit from MAPKi. Prospective clinical studies with novel MAPKi regimens are warranted in these patients.

**Highlights:**

- A meta-analysis describing clinical outcomes with MAPK targeted therapy in MAP2K1 mutant tumors.
- Clinical validation of MAP2K1 mutation Class as a predictive biomarker.
- Class 2 MAP2K1 mutations are sensitive to MEK-inhibitor containing regimens.

**Graphical Abstract:** 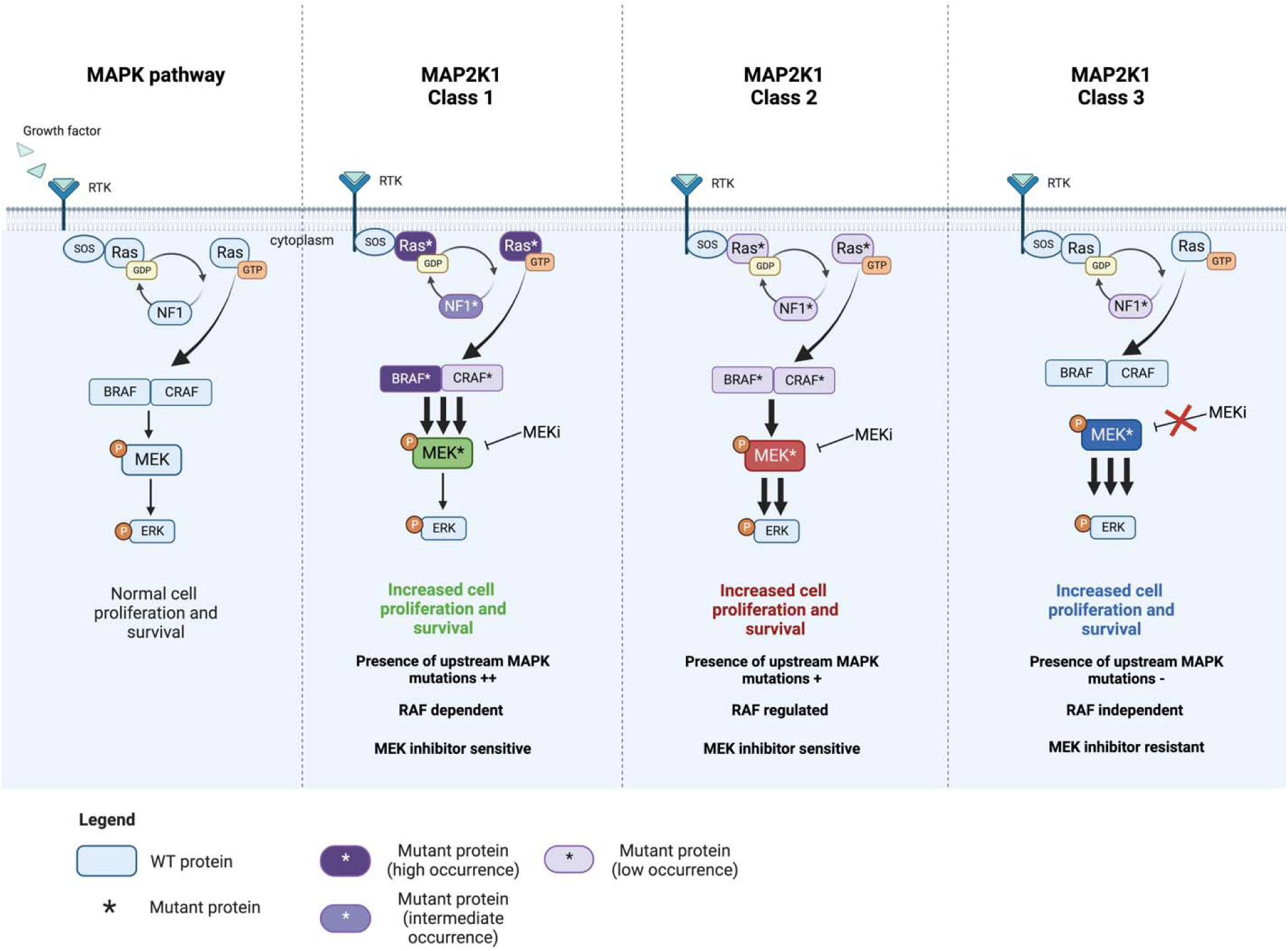

## Introduction

MAP2K1 (MEK1) is a kinase and a key constituent of the mitogen-activated protein kinase (MAPK) pathway which drives cellular proliferation and survival^1–3^. It is a recurrently mutated oncogene in human cancers, particularly Langerhans cell histiocytosis, melanoma, colorectal cancer, and non-small cell lung adenocarcinoma^4–8^.

Several hotspot driver mutations in MAP2K1 have been defined genetically and biochemically. These mutations were identified in treatment naïve tumors as well as in those resistant to MAPK targeted therapies^7,9–15^. A number of MAP2K1 mutations have been demonstrated to be oncogenic via their ability to transform non-cancerous cells, or by increasing the phosphorylation of downstream ERK^10,11,13,16–28^. In 2018, Gao et al. proposed a classification system for MAP2K1 mutations^2,13^. Class 1 MAP2K1 mutations are weak oncogenes which frequently co-occur with other MAPK pathway mutations. They are classified as RAF-dependent, as phosphorylation by RAF hyper-stimulates these mutants compared to wild-type MAP2K1^13^. Class 2 variants are considered to be RAF-regulated. They are still dependent on upstream RAF activity but to a lesser extent than class 1 mutations given that they can more potently activate the MAPK pathway without upstream RAF. Class 2 mutations occasionally, but not universally, co-occur with other MAPK pathway mutations^13^. Class 3 MAP2K1 mutations are unique in that they are entirely RAF-independent. These mutations are capable of autophosphorylation, and rarely co-occur with other MAPK pathway alterations^13^. Class 3 MAP2K1 mutations are also structurally distinct from wild-type, Class 1 and Class 2 MAP2K1 in such a way that impairs the ability of the FDA-approved MEK inhibitors cobimetinib, trametinib and binimetinib, from binding to these mutants^13^.

While this classification system proposes hypotheses for how tumors harboring MAP2K1 mutations of different classes may respond to MAPK targeted therapy, there is a paucity of clinical data to corroborate the preclinical data used to establish this paradigm. The totality of clinical evidence in this area is derived from case reports, case series, and individual patients from clinical trials whose tumors harbour a MAP2K1 mutation. When standard treatment options have been exhausted, many oncologists will provide off-label MAPK targeted therapies to these patients. Therefore, to establish a reference cohort to help guide treatment decisions and inform future clinical trial design, we synthesized all clinical evidence wherein patients with MAP2K1-mutant tumors were treated with MAPK targeted therapy. Our findings indicate that subsets of MAP2K1 mutant tumors can indeed be treated with currently available MAPK targeted therapies, particularly those with Class 2 but not Class 3 MAP2K1 mutations. These results provide important clinical insight into the activity of MAPK targeted therapies in tumors with MAP2K1 mutations and highlight groups of patients with MAP2K1 mutations where more effective targeted therapies are required.

## Methods

### AACR GENIE Analysis

Data on tumors with class 1, 2 and 3 MAP2K1 mutations was accessed from the AACR GENIE v13 dataset. To identify co-occurring mutations that were associated with MAP2K1 classes, we included only genes that were mutated in a minimum of 15 MAP2K1 mutant tumors, followed by filtering for p & q < 0.05 (p-value was obtained from a Chi-squared test and q-value from the Benjamini-Hochberg procedure). All frequently occurring MAPK pathway driver genes (NF1, NRAS, KRAS, BRAF, RAF1) and the top 10 most frequently altered non-MAPK genes were included in the oncoprint shown in Figure 1C.

**Figure 1:**
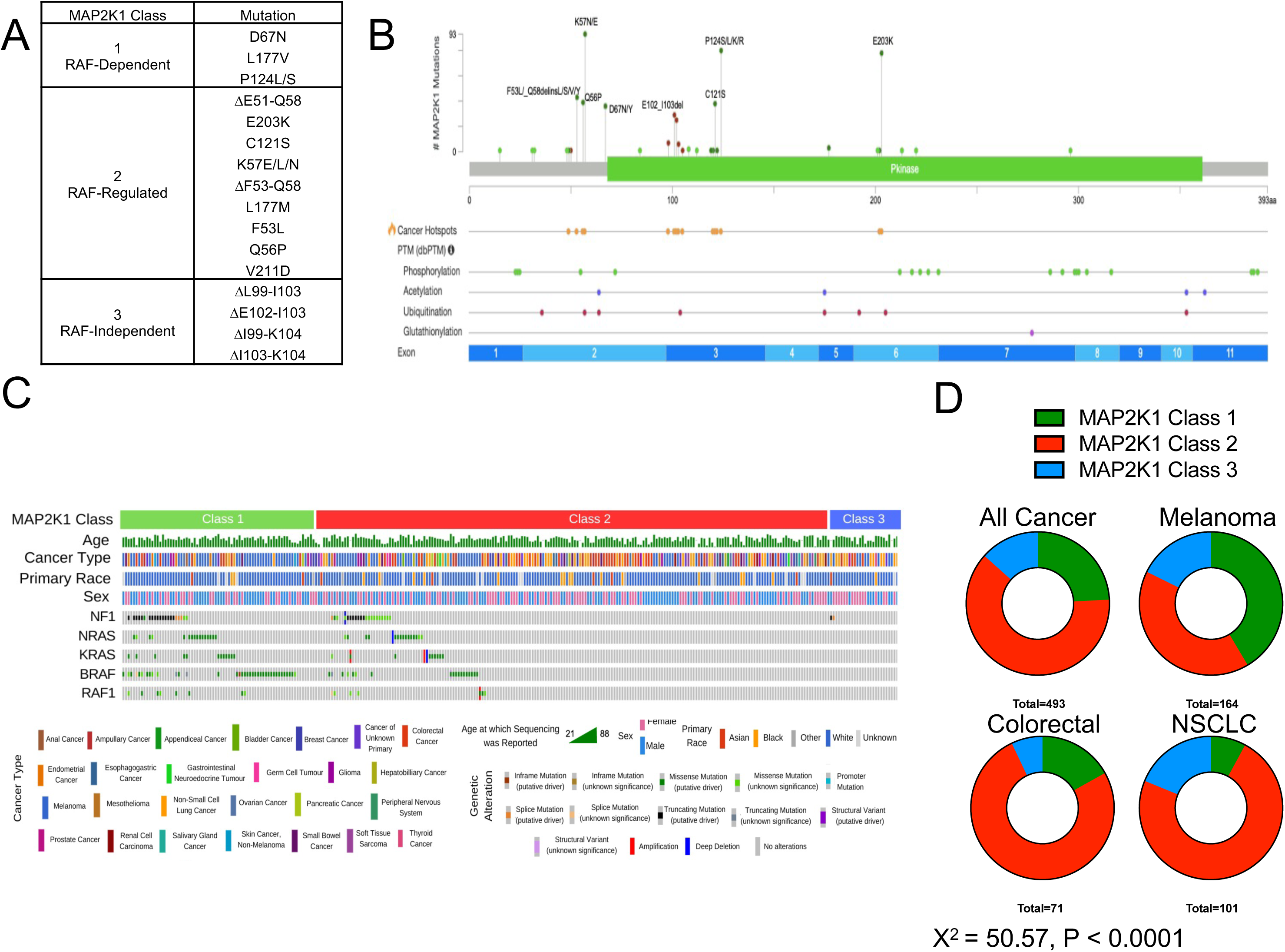
MAP2K1 mutation characteristics. A) Classification scheme of MAP2K1 mutations by Class. Derived from Gao *et al*. 2018 Cancer Discovery, and Gao *et al.* 2019 Cancer Discovery. B) Lollipop plot derived from all cancer types in AACR GENIE v13 demonstrating mutations in MAP2K1. C) Oncoprint derived from AACR GENIE v13 demonstrating co-occurring mutations in tumors harboring MAP2K1 mutations, separately by Class. For all genes selected, p<0.001 & q < 0.05 (p-value was obtained from a Chi-squared test and q-value from the Benjamini-Hochberg procedure). D) Breakdown of MAP2K1 mutation Class by primary cancer type in the AACR GENIE v13 dataset. P-value calculated by Chi-square test.

### Search Strategy

An extensive literature search was conducted from January 2010 to September 16 2022, in the following databases: Medline ALL (Medline and Medline Epub Ahead of print and In-Process & Other Non-Indexed Citations), Embase, Cochrane Central Register of Controlled Trials, Cochrane Database of Systematic Reviews, all from the OvidSP platform, and Web of Science from Clarivate Analytics. Where available, both controlled vocabulary terms and text words were used (Appendix 1). There were no language or study design restrictions. Published conference papers were included. The reference lists of included studies were scanned to identify additional relevant studies. The American Association for Cancer Research (AACR), the American Society of Clinical Oncology (ASCO) and the European Society of Medical Oncology (ESMO) were searched to identify any relevant conference papers. The study protocol was prospectively uploaded to PROSPERO (ID: CRD42022364061) and followed the preferred reporting items for systematic reviews and meta-analyses (PRISMA) guidelines^29^.

Abstracts were screened by two independent reviewers using Covidence software. Conflicts were resolved by a third reviewer. Response data and patient demographics were extracted by two independent reviewers. After data was extracted from all included publications, missing data was identified and requested from the original authors with two separate email prompts spaced out >7 days apart in the event of no reply.

### Inclusion and exclusion criteria

Inclusion criteria were: published reports of cancer patients with individual patient data describing 1) solid metastatic tumors, 2) harboring MAP2K1 mutations, 3) treatment with MAPK targeted therapies against EGFR, BRAF, MEK and/or ERK, and 4) availability of treatment response data. Exclusion criteria were: hematological cancers, pediatric cancers, adjuvant or neoadjuvant treatment setting.

### Primary and secondary outcomes

Primary outcome was progression free survival (PFS). Secondary outcomes were overall treatment response rate (ORR), duration of response (DOR) and overall survival (OS). When appropriate response criteria were used (Response evaluation criteria in solid tumors; RECIST1.1), patients with partial response or complete response were considered to have had a treatment response and those with stable disease or progressive disease were considered nonresponders^30^. When RECIST1.1 criteria were not used, response was recorded based on the primary paper’s author’s assessment of response or calculated from tumor measurements on computed tomography or magnetic resonance imaging provided in the text. For PFS analysis, patients were censored if there was no indication of progression or death at the time of last follow-up.

### Quality Assessment

To assess the methodological quality of individual studies included in the systematic review, we used a previously described tool that is adapted for evaluation of case reports and case series^31,32^. The tool includes 5 items that are derived from the Newcastle-Ottawa scale^33^. These 5 items examine the selection and representativeness of cases and the ascertainment of outcome and exposure, with each item scored one point if the information was specifically reported. We deemed the study to be of good quality (low risk of bias) when all 5 criteria were met, of moderate quality when 4 criteria were met, and of poor quality (high risk of bias) when ≤3 criteria were fulfilled.

### Statistical Analyses

We performed one-stage meta-analyses of pooled individual patient-level data from all included studies. Hazard ratio (HR) was used as the parameter of interest for PFS and OS, and odds ratio (OR) was used as the parameter of interest for response. A multi-level mixed-effects logistic regression model, incorporating individual study as a random effect, was used to estimate the ORs of responses between groups and its associated 95% confidence interval (95% CI). To analyze PFS, a shared frailty Cox regression model was used to account for heterogeneity across studies for all primary analyses. Multivariable Cox proportional hazards regression models were used to estimate adjusted HR (aHR), also with a multi-level mixed-effects Cox proportional hazards regression model that incorporated individual study as a random effect. All variables with P < 0.05 in univariable analysis were incorporated into the multivariable model. We tested the proportional hazards assumption by plotting the Schoenfeld residuals for each univariable and multivariable analysis, and they appeared random. Survival curves were visualized and evaluated with the Kaplan-Meier method and the log-rank test. Statistical analyses were performed with STATA v13 (StataCorp LLC, College Station, Texas, USA).

### Graphical Abstract

The graphical abstract was made with Biorender online software. The color coding indicating the low, intermediate or high incidence of co-occurring mutations is based on the incidence of the co-occurring alterations in the AACR Project GENIE v13 dataset.

## Results

### Characteristics of MAP2K1-mutated cancers in the AACR GENIE database

We analyzed the AACR GENIE database to identify patients whose tumors harbored MAP2K1 mutations^34^. Tumors with MAP2K1 mutations were grouped by class based on the existing classification scheme (Figure 1A)^10,13^. We identified a total of 493 tumors with MAP2K1 mutations, of which 119, 308, and 66, were class 1, 2, and 3, respectively (Figure 1B and C). We generated an oncoprint to visualize patient characteristics and co-mutations by MAP2K1 mutant class (Figure 1C). Consistent with previous reports^13^, we observed that the vast majority of tumors with Class 1 MAP2K1 mutations (82.35%) harbor at least one concurrent MAPK pathway driver mutation (in one or more of NF1, NRAS, KRAS, BRAF, RAF1), while Class 2 (32.79%) and Class 3 (9.09%) mutant tumors are less likely to have co-existing MAPK pathway driver mutations (Figure 1C). We observed significant differences in the distribution of MAP2K1 mutation class by primary tumor type, with melanomas having a higher prevalence of Class 1 MAP2K1 mutations compared to colorectal and lung cancer (P<0.0001) (Figure 1D).

### Characteristics of included studies and patients in meta-analysis

We identified 81,701 articles in our search (Supplemental Figure S1). After removing ineligible articles, a total of 20 articles were included in the meta-analysis (Appendix 2), comprising a total of 46 patients with MAP2K1 mutant-tumors and who were treated with MAPK targeted therapy (MAPK TT) in the metastatic setting (Supplemental Figure S1). 13 patients received concurrent chemotherapy with MAPK TT (Table 1). A more detailed description of the different MAPK TT regimens used for patients in the study is presented in Supplemental Table S1 and included approved inhibitors of BRAF, MEK, and EGFR. Of 26 patients with previous treatment data available, 11 patients received MAPK TT in the first-line metastatic setting, and 15 patients had received prior lines of therapy (Table 1). We also performed risk-of-bias (ROB) assessment for all studies included in the meta-analysis on a five-point scale (Supplemental Figure S2).

**Table 1:** Individual patient characteristics.

|  | Entire Cohort | Unclassified | Class 1 | Class 2 | Class 3 | Fischer's Exact Test P-Value | Pearson's Chi-Squared |
| --- | --- | --- | --- | --- | --- | --- | --- |
| *Entire Cohort* | 46 | 3 | 15 | 26 | 2 |  |  |
| *Study Characteristics* |  |  |  |  |  |  |  |
| **Study Type** |  |  |  |  |  |  |  |
| Prospective | 21 | 0 | 11 | 10 | 0 | **0.015** |  |
| Retrospective | 25 | 3 | 4 | 16 | 2 |  |  |
| **Response Criteria** |  |  |  |  |  |  |  |
| RECIST | 32 | 2 | 14 | 16 | 0 | **0.012** |  |
| non-RECIST | 14 | 1 | 1 | 10 | 2 |  |  |
| **Geographic Location** |  |  |  |  |  |  |  |
| North America | 37 | 3 | 13 | 19 | 2 | 0.676 |  |
| Other | 9 | 0 | 2 | 7 | 0 |  |  |
| *Patient & Treatment Characteristics* |  |  |  |  |  |  |  |
| **Gender** |  |  |  |  |  |  |  |
| Male | 21 | 2 | 6 | 13 | 0 | 0.278 |  |
| Female | 20 | 1 | 9 | 8 | 2 |  |  |
| Unknown | 5 |  |  |  |  |  |  |
| **Age** |  |  |  |  |  |  |  |
| >= 65 | 12 | 0 | 5 | 7 | 0 | 1 |  |
| < 65 | 27 | 2 | 10 | 13 | 2 |  |  |
| Unknown | 7 |  |  |  |  |  |  |
| **Cancer Type** |  |  |  |  |  |  |  |
| Melanoma | 19 | 2 | 14 | 3 | 0 | **<0.001** | **34.797; P < 0.001** |
| Colorectal Cancer | 12 | 1 | 0 | 9 | 2 | **0.002** |  |
| Lung Cancer | 11 | 0 | 0 | 11 | 0 | **0.006** |  |
| Other | 4 | 0 | 1 | 3 | 0 | 1.000 |  |
| **Therapy Type** |  |  |  |  |  |  |  |
| MEKi-containing regimen | 24 | 2 | 5 | 16 | 1 | 0.304 |  |
| Non-MEKi-containing regimen | 22 | 1 | 10 | 10 | 1 |  |  |
| **Concomitant Chemotherapy** |  |  |  |  |  |  |  |
| Yes | 13 | 0 | 0 | 12 | 1 | **0.002** |  |
| No | 33 | 3 | 15 | 14 | 1 |  |  |
| **Prior Lines of Therapy** |  |  |  |  |  |  |  |
| 0 | 11 | 0 | 0 | 10 | 1 | 0.742 | 0.477 |
| 1+ | 15 | 2 | 1 | 11 | 15 |  |  |
| **Co-Occurring BRAF V600 Mutation** |  |  |  |  |  |  |  |
| Present | 18 | 1 | 14 | 3 | 0 | **<0.001** |  |
| Absent | 28 | 2 | 1 | 23 | 2 |  |  |
| **Co-Occurring MAPK Mutation** |  |  |  |  |  |  |  |
| Present | 25 | 1 | 15 | 9 | 0 | **<0.001** |  |
| Absent | 21 | 2 | 0 | 17 | 2 |  |  |
| **Co-Occurring non-MAPK Oncogenic Mutation** |  |  |  |  |  |  |  |
| Present | 17 | 1 | 4 | 10 | 2 | 0.261 |  |
| Absent | 29 | 2 | 11 | 16 | 0 |  |  |

Among the 46 patients included in the meta-analysis, 15 harbored Class 1 MAP2K1 mutations, 26 had Class 2 mutations, 2 had Class 3 mutations, and 3 had MAP2K1 mutations yet to be classified (Table 1). Patients with Class 1 mutant tumors were more likely to derive from prospective studies using RECIST criteria to define treatment response, be of melanoma primary harboring concurrent BRAF mutations and be treated with BRAF inhibitors (Table 1). Patients with Class 2 MAP2K1 mutations were most likely to be treated with concurrent chemotherapy in addition to MAPK TT (Table 1).

### Characteristics associated with response to MAPK TT in MAP2K1 mutant tumors

In the entire population, 13 out of 46 patients (28%) experienced a treatment response (Figure 2A). There were no patient characteristics that predicted treatment response globally with statistical significance (Supplemental Table S2). However, we observed an increased response rate in patients whose tumors harbour Class 2 MAP2K1 mutations (7/16 responded) versus tumors with other MAP2K1 mutations (0/8 responded) when treated with regimens including MEK inhibitors (P=0.05) (Figure 2B). Indeed, 6/7 patients with Class 2 MAP2K1 mutations that responded to MAPK targeted therapies had received a MEK inhibitor as part of their treatment regimen. Patients experienced similar response rates regardless of whether they had concurrent additional MAPK pathway mutations (Figure 2C).

**Figure 2:**
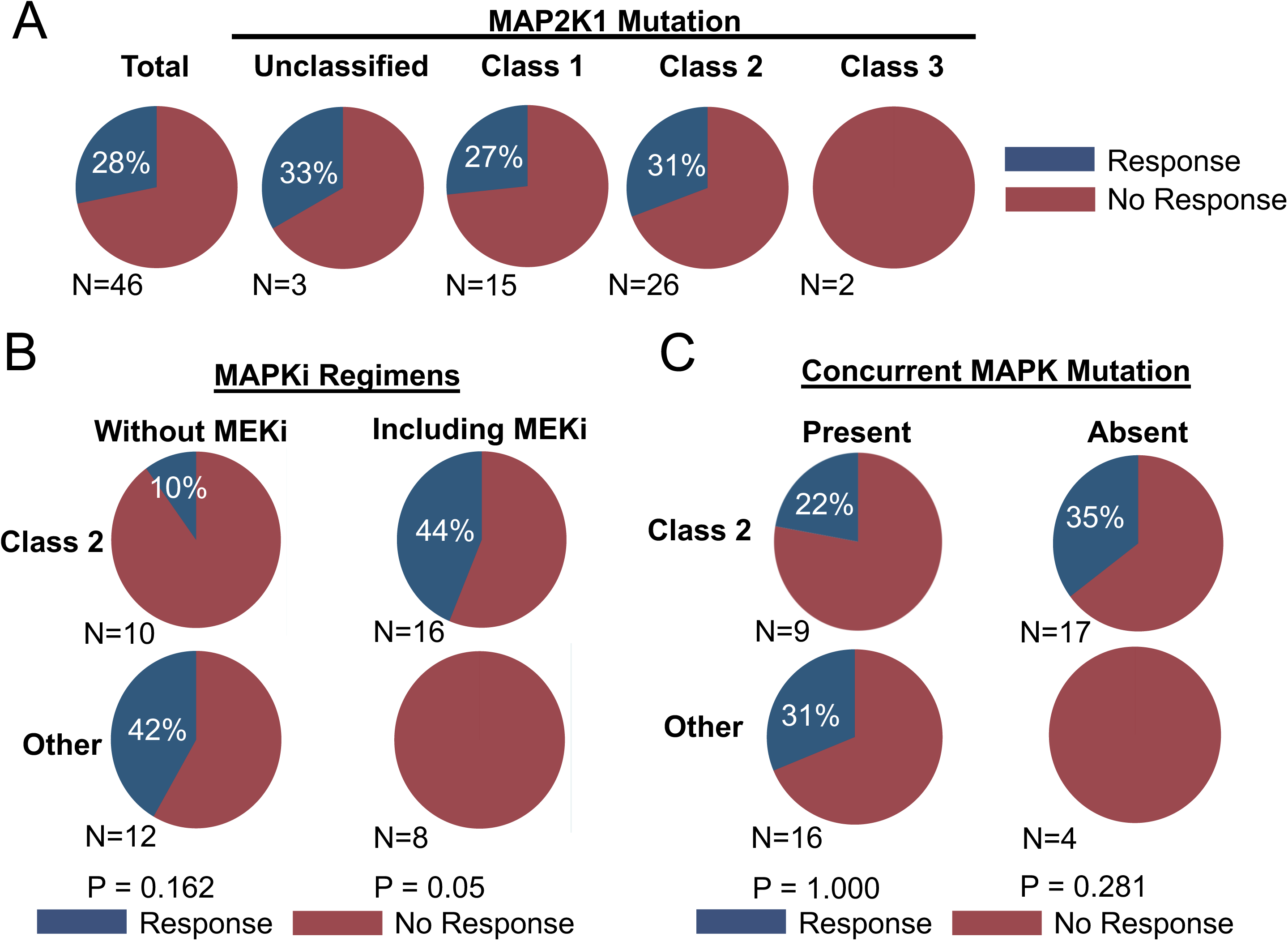
A) Overall response rates in the whole cohort (total) according to MAP2K1 mutant class. B) Comparison of overall response rate between Class 2 versus other MAP2K1 mutations depending on whether MEK inhibitors were used as part of the treatment regimen. C) Comparison between overall response rates in patients with Class 2 versus other MAP2K1 mutations treated with MAPK targeted therapy based upon the presence or absence of concurrent MAPK pathway oncogenic mutations. The MAPK pathway driver mutations identified in the treated patient cohort were found in BRAF, NRAS, KRAS and EGFR. P-values calculated with Fischer’s Exact Test.

### Characteristics associated with progression-free and overall survival in MAP2K1 mutant tumors treated with MAPK TT

In the entire population, median PFS was 3.9 months (Table 2), and median OS was 15 months (Table 3). Patients with Class 2 MAP2K1 mutations experienced prolonged PFS (median PFS 5 months, P=0.04) and OS (median not reached, P=0.0001) compared to the rest of the cohort (Figure 3A and B), while patients with unclassified MAP2K1 mutations experienced the shortest PFS (median 1.2 months, HR: 4.2, 95% CI: 1.1-15.6, P=0.035) (Table 2). Class 2 mutant tumors treated with MEK inhibitor-containing regimens trended towards prolonged PFS (P=0.06) and experienced longer OS (P=0.0056) (Figure 3C and D).

**Figure 3:**
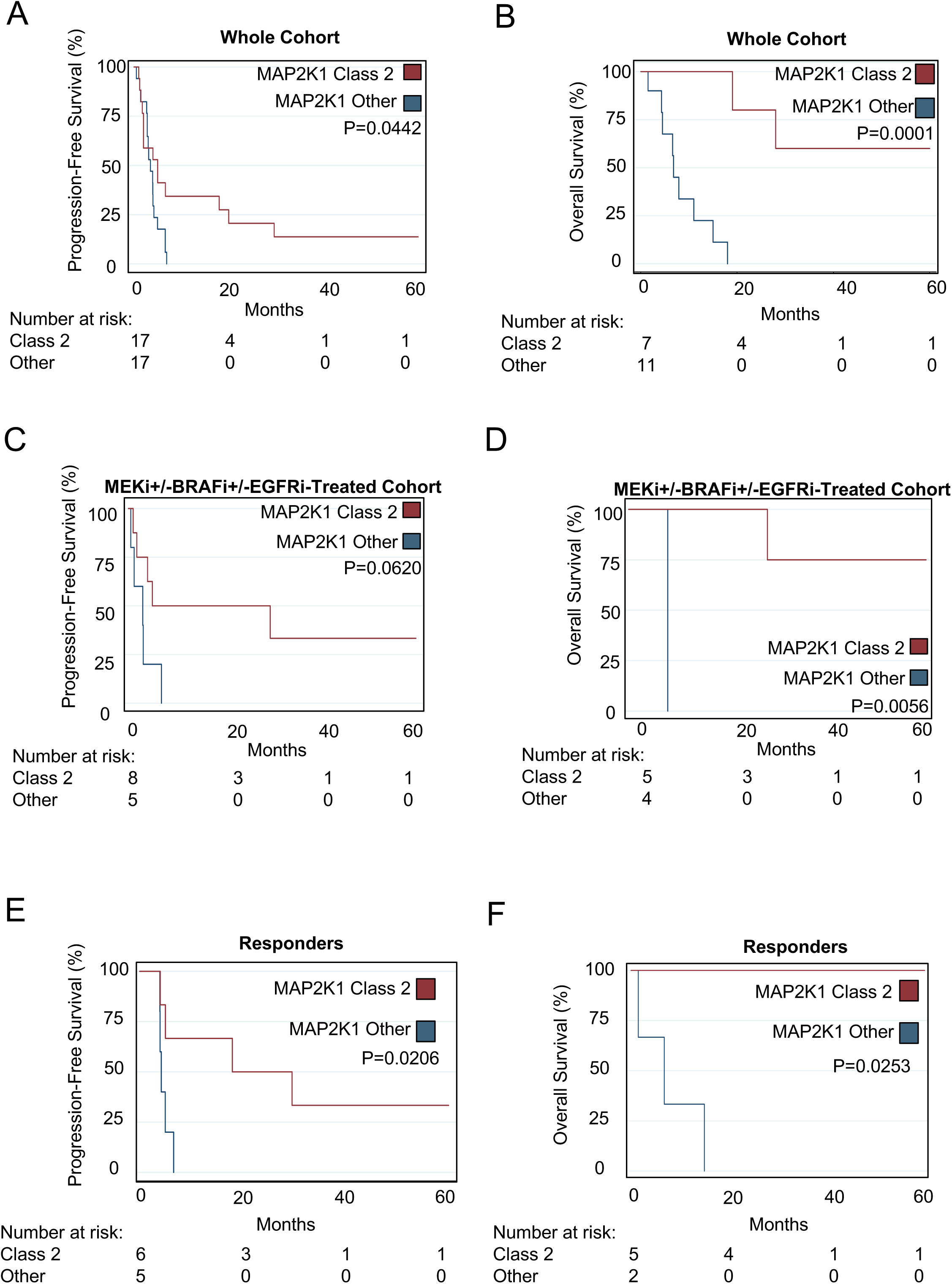
Relationship between MAP2K1 Class, clinical features, and survival. A) Progression-free survival and B) overall survival in Class 2 versus other MAP2K1 mutant tumors in the whole MAPK targeted therapy-treated cohort. C) Progression-free survival and D) overall survival in Class 2 versus other MAP2K1 mutant tumors in patients treated with MEK inhibitor-containing treatment regimens. E) Progression-free survival and F) overall survival in patients who experienced a response to MAPK targeted therapy. P-values calculated with log-rank test.

**Table 2:** Progression-free survival associated with clinical variables. NR: Not reached

|  | # of Patients | # of Patients with PFS Data | Median PFS | Hazard Ratio | 95% Confidence Interval | P-Value | Adjusted Hazard Ratio | Adjusted 95% Confidence Interval | Adjusted P-Value |
| --- | --- | --- | --- | --- | --- | --- | --- | --- | --- |
| Entire Cohort | 46 | 35 | 3.9 |  |  |  |  |  |  |
| Study Characteristics |  |  |  |  |  |  |  |  |  |
| Study Type |  |  |  |  |  |  |  |  |  |
| Prospective | 21 | 10 | 3.1 | 1.214 | 0.487 - 3.027 | 0.677 |  |  |  |
| Retrospective | 25 | 24 | 3.9 |  |  |  |  |  |  |
| Response Criteria |  |  |  |  |  |  |  |  |  |
| RECIST | 32 | 22 | 3.5 | 1.084 | 0.428 - 2.743 | 0.865 |  |  |  |
| non-RECIST | 14 | 12 | 4 |  |  |  |  |  |  |
| Geographic Location |  |  |  |  |  |  |  |  |  |
| North America | 37 | 27 | 3.5 | 2.331 | 0.887 - 6.121 | 0.086 |  |  |  |
| Other | 9 | 7 | 6.6 |  |  |  |  |  |  |
| Patient & Treatment Characteristics |  |  |  |  |  |  |  |  |  |
| Gender |  |  |  |  |  |  |  |  |  |
| Male | 21 | 14 | 3.1 | 1.15 | 0.449 - 2.949 | 0.771 |  |  |  |
| Female | 20 | 15 | 4 |  |  |  |  |  |  |
| Age |  |  |  |  |  |  |  |  |  |
| >= 65 | 12 | 8 | 2 | 1.784 | 0.652 - 4.880 | 0.259 |  |  |  |
| < 65 | 27 | 19 | 4 |  |  |  |  |  |  |
| Cancer Type |  |  |  |  |  |  |  |  |  |
| Melanoma | 19 | 15 | 3.9 | 1.415 | 0.686 - 2.919 | 0.347 |  |  |  |
| Colorectal Cancer | 12 | 11 | 3 | 1.659 | 0.781 - 3.525 | 0.188 |  |  |  |
| Lung Cancer | 11 | 4 | 1.5 | 2.455 | 0.514 - 11.722 | 0.260 |  |  |  |
| Other | 4 | 4 | NR | 0.0835 | 0.0111 - 0.627 | **0.016** | 0.0881 | 0.0116 - 0.668 | **0.019** |
| Therapy Type |  |  |  |  |  |  |  |  |  |
| MEKi-containing regimen | 24 | 13 | 4 | 0.55 | 0.248 - 1.221 | 0.142 |  |  |  |
| Non-MEKi-containing regimen | 22 | 21 | 3.9 |  |  |  |  |  |  |
| Concomitant Chemotherapy |  |  |  |  |  |  |  |  |  |
| Yes | 13 | 6 | 4 | 0.514 | 0.150 - 1.762 | 0.29 |  |  |  |
| No | 33 | 28 | 3.5 |  |  |  |  |  |  |
| Prior Lines of Therapy |  |  |  |  |  |  |  |  |  |
| 0 | 11 | 6 | 6.6 | 0.773 | 0.773 - 12.681 | 0.11 |  |  |  |
| 1+ | 15 | 11 | 2 |  |  |  |  |  |  |
| Co-Occurring MAPK Mutation |  |  |  |  |  |  |  |  |  |
| Present | 25 | 14 | 3.9 | 1.066 | 0.461 - 2.460 | 0.882 |  |  |  |
| Absent | 21 | 20 | 3 |  |  |  |  |  |  |
| Co-Occurring BRAF V600 Mutation |  |  |  |  |  |  |  |  |  |
| Present | 18 | 14 | 3.9 | 1.065 | 0.461 - 2.460 | 0.882 |  |  |  |
| Absent | 28 | 20 | 3 |  |  |  |  |  |  |
| Co-Occurring non-MAPK Oncogenic Mutation |  |  |  |  |  |  |  |  |  |
| Present | 17 | 11 | 4 | 0.818 | 0.319 - 2.099 | 0.677 |  |  |  |
| Absent | 29 | 23 | 3.9 |  |  |  |  |  |  |
| MAP2K1 Class |  |  |  |  |  |  |  |  |  |
| Unclassified | 3 | 3 | 1.2 | 4.160 | 1.109 - 15.609 | **0.035** | 2.627 | 0.730 - 9.449 | 0.139 |
| 1 | 15 | 12 | 3.5 | 1.422 | 0.666 - 3.041 | 0.363 |  |  |  |
| 2 | 26 | 17 | 5 | 0.456 | 0.208 - 1.002 | **0.051** | 0.531 | 0.226 - 1.244 | 0.145 |
| 3 | 2 | 3 | 3 | 1.360 | 0.282 - 6.537 | 0.701 |  |  |  |

**Table 3:** Overall survival associated with clinical variables. *NR: Not Reached NA: Unable to be calculated

|  | # of Patients | # of Patients with OS Data | Median OS | Hazard Ratio | 95% Confidence Interval | P-Value |
| --- | --- | --- | --- | --- | --- | --- |
| *Entire Cohort* | 46 | 18 | 15 |  |  |  |
| *Study Characteristics* |  |  |  |  |  |  |
| **Study Type** |  |  |  |  |  |  |
| Prospective | 21 | 10 | 6.8 | 26.061 | 2.481 - 273.660 | **0.007** |
| Retrospective | 25 | 8 | NR |  |  |  |
| **Response Criteria** |  |  |  |  |  |  |
| RECIST | 32 | 13 | 11 | 0.661 | 0.0260 - 16.805 | 0.802 |
| non-RECIST | 14 | 5 | 19 |  |  |  |
| **Geographic Location** |  |  |  |  |  |  |
| North America | 37 | 12 | 11 | 0.665 | 0.0445 - 9.928 | 0.767 |
| Other | 9 | 6 | 19 |  |  |  |
| *Patient & Treatment Characteristics* |  |  |  |  |  |  |
| **Gender** |  |  |  |  |  |  |
| Male | 21 | 8 | NR | 0.231 | 0.0264 - 2.012 | 0.184 |
| Female | 20 | 10 | 7.9 |  |  |  |
| **Age** |  |  |  |  |  |  |
| ≥/ 65 | 12 | 3 | 18 | 1.485 | 0.205 - 10.768 | 0.696 |
| < 65 | 27 | 13 | 6.6 |  |  |  |
| **Cancer Type** |  |  |  |  |  |  |
| Melanoma | 19 | 11 | 6.8 | 31.811 | 0.842 - 1202.314 | 0.062 |
| Colorectal Cancer | 12 | 3 | 19 | 0.199 | 0.00554 - 7.176 | 0.378 |
| Lung Cancer | 11 | 0 | NA | NA | NA | NA |
| Other | 4 | 4 | NR | 0.110 | 0.00767 - 1.574 | 0.104 |
| **Therapy Type** |  |  |  |  |  |  |
| Non-MEKi-containing regimen | 22 | 9 | 7.9 | 0.291 | 0.0680 - 1.242 | 0.095 |
| MEKi-containing regimen | 24 | 9 | NR |  |  |  |
| **Concomitant Chemotherapy** |  |  |  |  |  |  |
| Yes | 13 | 3 | 19 | 0.199 | 0.006 - 7.187 | 0.378 |
| No | 33 | 15 | 11 |  |  |  |
| **Prior Lines of Therapy** |  |  |  |  |  |  |
| 0 | 11 | 5 | 19 | 0.456 | 0.0469 - 4.447 | 0.500 |
| 1+ | 15 | 5 | NR |  |  |  |
| **Co-Occurring MAPK Mutation** |  |  |  |  |  |  |
| Present | 25 | 10 | 6.8 | 58.072 | 1.509 - 2234.224 | **0.029** |
| Absent | 21 | 8 | 28 |  |  |  |
| **Co-Occurring BRAF Mutation** |  |  |  |  |  |  |
| Present | 18 | 10 | 6.8 | 58.072 | 1.509 - 2234.224 | **0.029** |
| Absent | 24 | 8 | 28 |  |  |  |
| **Co-Occurring non-MAPK Oncogenic Mutation** |  |  |  |  |  |  |
| Present | 17 | 6 | 18 | 0.476 | 0.030 - 7.460 | 0.597 |
| Absent | 29 | 12 | 11 |  |  |  |
| **MAP2K1 Class** |  |  |  |  |  |  |
| Unclassified | 3 | 1 | NA | NA | NA | NA |
| 1 | 15 | 9 | 6.8 | NA | NA | NA |
| 2 | 26 | 7 | NR | NA | NA | NA |
| 3 | 2 | 1 | NA | NA | NA | NA |

Patients with non-melanoma/lung/colorectal primaries experienced longer PFS but not OS compared to the rest of the cohort (median PFS not reached; HR: 0.08, 95% CI: 0.01–0.63, P=0.02 and median OS not reached; HR: 0.1, 95% CI: 0.008-1.6, P=0.1) (Table 2, Table 3, Supplemental Figure 3A and B). The same trends were observed when patients were treated specifically with MEK inhibitor-containing regimens (PFS P=0.01, OS P=0.07; Supplemental Figure 3C and D). In multivariable analyses, non-melanoma/lung/colorectal primaries remained independently associated with prolonged PFS (aHR 0.09, 95% CI: 0.012 – 0.67, P=0.019).

Amongst patients who responded to MAPK targeted therapies, those with Class 2 MAP2K1 mutations experienced prolonged duration of response (P=0.02) and overall survival (P=0.03) compared to those with other MAP2K1 mutations (Figure 3E and F). Depth of response, stratified by RECIST categories of complete response, partial response, stable disease, or progression disease, was associated with both PFS and OS (Supplemental Figure S4A and B).

### Quality assessment

The majority of the patients included in this analysis were reported in retrospective studies (25/46; 54%). These may be more subject to bias than prospective studies. Indeed, we observed prolonged OS but not PFS or ORR in patients derived from retrospective studies (Table 2, Table 3, Supplemental Table S2). To better characterize ROB and its impact on our results, we performed a quality assessment of all included studies, using a validated five-point scale (Supplemental Figure S2). We analyzed whether ROB among the studies was associated with treatment response. There was no statistically significant difference in RR (22% versus 35%, P=0.5) between patients derived from studies with high ROB (score 0-3) compared with those with low/moderate ROB (score 4-5) (Supplemental Figure S5A). Similarly, we observe no differences in PFS (P=0.6) or OS (P=0.1) when patients are stratified by ROB (Supplemental Figure S5B and C).

## Discussion

Oncogenic MAP2K1 mutations are identified in approximately 5% of melanomas and 1-2% of lung and colorectal cancers^34^. Herein, we have assembled the largest clinical cohort of patients with MAP2K1 mutant tumors with associated treatment response to date. This has allowed us to perform comprehensive analyses of characteristics associated with response to MAPK TT in this population. Our results highlight the importance of testing for the presence of MAP2K1 mutations in patients with many types of advanced cancer. This goes beyond the most common cancer types with MAP2K1 mutations (melanoma, colorectal, lung) given that we observed prolonged PFS in univariable and multivariable analyses in metastatic cancers with MAP2K1 mutations treated with MAPK TT that are non-melanoma/colorectal/lung primary. In our cohort, these non-melanoma/colorectal/lung primary tumors included ovarian, skin squamous cell carcinoma, cholangiocarcinoma, and spindle cell neoplasm^11,35–37^.

We observed similar outcomes in patients with colorectal and non-colorectal tumor types. This contrasts from existing data for other MAPK targeted therapt contexts, such as BRAF inhibitors for BRAF V600 mutant cancers, where colorectal cancer patients tend to experience inferior outcomes compared to lung cancer and melanoma patients^38^. This can be explained by the fact that of the 11/12 colorectal cancer patients in our cohort received EGFR inhibitor-containing regimens, overcoming the known intrinsic resistance mechanism of EGFR mediated feedback in colorectal cancers treated with MAPK pathway inhibitors^39,40^.

In addition to their role as *de novo* driver mutations, MAP2K1 mutations have also been demonstrated to emerge in the context of resistance to EGFR, BRAF and MEK MAPK TT^10,21,41–44^. This suggests that the frequency of these mutations in the treatment-refractory setting may be higher than in treatment naïve tumors. Recently, MAP2K1 mutations have emerged as important resistance mechanisms in cancers treated with the KRAS G12C inhibitor sotorasib. Indeed, 4/6 colorectal cancers treated with sotorasib acquired *de novo* MAP2K1 mutations upon resistance^15^. It remains to be seen whether similar rates of MAP2K1 mutations will be seen in larger populations treated with this novel class of targeted therapies. However, this finding underscores the importance of better understanding the therapeutic vulnerabilities of cancers with MAP2K1 alterations.

Our findings in human patients largely validate the classification scheme established in the preclinical literature^10,13^. Namely, Class 1 MAP2K1 mutations may be sensitive to MAPK TT, however only when the universally co-occurring MAPK pathway mutational driver is also targeted. This is highlighted by the fact that in our dataset, all 4 Class 1 MAP2K1 mutant tumors that experienced a response to treatment were in the context of co-existing BRAF V600E mutations treated with BRAF inhibitors.

In contrast to Class 1 MAP2K1 mutations that are weakly oncogenic, Class 3 mutations have been reported to be potent oncogenes that rarely co-occur with concurrent MAPK driver mutations^13^. However, Class 3 MAP2K1 mutations result in a conformational change in the protein structure that impairs the ability of MEK inhibitors to bind these mutants^13^. Our findings, albeit limited by small sample size, corroborate the preclinical data, with 2/2 Class 3 mutant tumors in our dataset experiencing progressive disease on treatment with panitumumab and trametinib, respectively. This underscores a need for novel therapeutic strategies for Class 3 MAP2K1 mutations – although this Class of MAP2K1 mutations is rare. This may include novel MEK inhibitors capable of inhibiting these mutant proteins^13^, or targeting downstream components of the MAPK signaling pathway, such as ERK^45,46^.

Class 2 MAP2K1 mutants fall between Class 1 and Class 3, in that they are more potent oncogenes that Class 1 but less so than Class 3. Class 2 MAP2K1 mutants occasionally but not universally co-occur with other MAPK driver mutations and can be effectively inhibited with FDA-approved MEK inhibitors^13^. The data presented herein suggests that patients whose tumors harbor Class 2 MAP2K1 mutations may be effectively treated with MAPK TT in clinical settings. MEK inhibitor-containing TT regimens, regardless of the status of other MAPK or non-MAPK driver oncogenes are compelling strategies for patients whose tumors harbor these mutations. To this effect, we observe prolonged PFS and OS in Class 2 mutant tumors compared to the rest of the cohort. Together, these findings nominate Class 2 MAP2K1 mutations as potentially targetable oncogenic mutations that should be considered for MAPK TT targeted therapy approaches in clinical trials and molecular tumor boards.

In our cohort we included all patients with MAP2K1 mutated tumors who were treated with FDA-approved MAPK TT, including two patients with unclassified MAP2K1 mutations (K59_60insEQK and I111S). Mutations at both of these amino acid residues have demonstrated high and moderate transforming capacity^16^, respectively, suggesting that these mutations are indeed oncogenic. This emphasizes the importance of continuing development of the existing classification scheme to include additional mutations.

Our findings are limited by both the small sample size of patients eligible for inclusion in our study, as well as the retrospective nature of many of the patients included in the meta-analysis. In performing risk of bias analysis, we partially mitigate this limitation. However, we do observe prolonged OS, but not ORR or PFS, in patients from retrospective compared to prospective studies. As such the results reported here may over-represent the expected outcomes experienced in real-world settings. However, the signals of clinical activity, especially with MEK inhibitors in Class 2 mutant tumors, support the development of prospective trials for these patients.

Finally, we are missing data on performance status, degree of tumor burden, and line of therapy for some patients, all of which may be important confounders to our results.

The rarity and variable oncogenicity of each MAP2K1 mutation remains a challenge for drug developers and may complicate interpretation of results from future prospective trials that may be developed for patients with these mutations. To facilitate effective drug development targeted against these important driver mutations, it will be critical for the community to collaborate in multicenter trials and share data regarding patient responses, tumor types, and co-mutation status whenever possible.

Taken together, the existing literature confirms many of the predictions presented by preclinical research with respect to differences between MAP2K1 mutant classes and establishes new hypotheses worthy of further investigation. Currently available MAPK TTs have demonstrated clinical activity in a subset of tumors with MAP2K1 mutations—especially those with Class 1 and 2 MAP2K1 mutations. However, to date, these MAPK-directed therapies appear to be associated with lower ORR than has been observed in patients with other MAPK drivers, such as BRAF V600E or EGFR^47,48^. Additionally, future studies may yield more benefit if therapeutic approaches are tailored according to MAP2K1 mutant Class and primary tumor type. Finally, because of modest ORRs with MAPK inhibitor monotherapies, future clinical trials should incorporate combination therapy strategies to target tumors with MAP2K1 mutations more effectively. This study provides a comprehensive analysis of the clinical outcomes of patients with MAP2K1 mutated tumors who received treatment with MAPK pathway inhibitors. We believe that this analysis will be useful for molecular tumor boards who provide treatment recommendations for patients with rare oncogenic alterations. Moreover, these data can be used to optimize the design of prospective clinical trials for patients with MAP2K1 mutant tumors.

## Data Availability

All data produced in the present study are available upon reasonable request to the authors

## Acknowledgments

ER acknowledges a Marathon of Hope Cancer Centers Network Health Informatics and Data Science Award. AANR acknowledges salary support from a Fonds de recherche du Québec – Santé (FRQS) Chercheuses-Boursières Cliniciennes Award. This research was supported by funding from the TransMedTech Institute and Apogee Canada Research Excellence Fund, and a Conquer Cancer Foundation of ASCO Career Development Award to AANR.

## Figure & Table Legends

**Supplemental Table S1:** MAPK targeted therapy regimens used for patients in the meta-analysis.

**Supplemental Table S2:** Overall Response Rates Associated With Clinical Variables.

**Supplemental Figure S1:** Preferred Reporting Items for Systematic Reviews and Meta-Analyses diagram demonstrating search and inclusion of studies for meta-analysis. diagram and response to treatment.

**Supplemental Figure S2**: Risk of bias assessment of the individual studies included in the systematic review. The 5-point score is adapted from the Newcastle-Ottawa score: 1) Selection – Did the patients represent all/consecutive patients with MAP2K1 mutations from the medical center? 2) Ascertainment (Diagnosis) – Was the diagnosis correctly made with pathology-proven cancer and next-generation sequencing assay to confirm MAP2K1 mutation? 3) Ascertainment (Outcome) – Was treatment response adequately ascertained using RECIST criteria? 4) Follow-up – Was follow up long enough for treatment responses to be evaluated? 5) Reporting – Is the case described with sufficient details (*e.g.,* drug posology, previous lines of chemotherapy) to allow other investigators to replicate the research or to allow practitioners make inferences related to their own practice? The 5 first columns represent the 5 assessment criteria (black circle = yes, grey circle = no). The total risk of bias score is the last column and the colors indicate the score.

**Supplemental Figure S3:** Relationship between primary tumor type survival. A) Progression-free survival and B) overall survival. C) Progression-free survival and D) overall survival stratified by primary tumor type exclusively in patients treated with MEK inhibitor containing regimens. P-values calculated with log-rank test.

**Supplemental Figure S4:** Progression-free survival (A) and overall survival (B) in patients included in the meta-analysis based upon response type. CR: complete response, PR: partial response, SD: stable disease, PD: progressive disease. P-values calculated with log-rank test.

**Supplemental Figure S5:** Outcome comparisons of patients from studies with high versus low/moderate risk of bias (ROB). A) Overall response rate in patients from studies with low/moderate (ROB score 4-5) versus high (ROB score 0-3) risk of bias. B) Progression-free survival in patients from studies with low/moderate (ROB score 4-5) versus high (ROB score 0-3) risk of bias. C) Overall survival in patients from studies with low/moderate (ROB score 4-5) versus high (ROB score 0-3) risk of bias. P-values calculated with Fischer’s Exact Test in A, and with log-rank test in B and C.

**Appendix 1**: Detailed search strategy.

**Appendix 2:** List of references of studies used to extract data in the meta-analysis.

## Author Contributions

Conceptualization: MD, AANR

Data curation: MD, ER, SP, FF, AN, AML, CVR, AS, DB, PT, AB, ML, AQ, NN, MA, AE, DBJ, ML, AANR.

Formal analysis: MD, ER, AANR

Methodology: MD, ER, AQ, AANR

Project administration: MD, AANR

Supervision: MD, NN, AE, DBJ, ML, AANR

Funding: AANR

Writing- original draft: MD, ER, AANR

Writing- review & editing: MD, ER, SP, FF, AN, AML, CVR, AS, DB, PT, AB, ML, AQ, NN, MA, AE, DBJ, ML, AANR.

