## Appendix 1 for "Clinical activity of Mitogen-Activated Protein Kinase (MAPK) inhibitors in patients with MAP2K1 (MEK1)-mutated metastatic cancers"

**Search documentation**

| Total with duplicates | 105 445 |
| --- | --- |
| Duplicates removed |  |
| Total without duplicates |  |

| Database | Ovid MEDLINE(R) ALL |
| --- | --- |
| Database time coverage | 1946-present |
| Date searched | 16 September 2022 |
| Total | 38 759 |
| Duplicates |  |

| Database | Ovid Embase |
| --- | --- |
| Database time coverage | 1996-present |
| Date searched | 16 September 2022 |
| Total | 66 686 |
| Duplicates |  |

**Search summary**

[Ovid MEDLINE(R) ALL <1946 to September 15, 2022>](https://proxy.library.mcgill.ca/login?url=http://ovidsp.ovid.com/ovidweb.cgi?T=JS&NEWS=N&PAGE=main&SHAREDSEARCHID=19YOh4Y3igdG0FgvI51EMbfjnepGQeUKfrRX017J5YGhdLdkH5TtaDVqh29LAXlod)

1	Proto-Oncogene Proteins c-raf/	3814

2	(craf or c raf).mp,kw.	4863

3	MAP Kinase Kinase 1/	3285

4	(mitogen activated protein kinase kinase 1 or mitogen activated protein kinase kinase one or map kinase kinase 1 or map kinase kinase one or map2k1 or mek1 or mek 1 or mkk1 or mkk 1 or erk1 or erk 1 or "mek1/2").mp,kw.	43800

5	MAP Kinase Kinase 2/	1150

6	(mitogen activated protein kinase kinase 2 or mitogen activated protein kinase kinase two or kinase activator kinase or mek2 or map kinase kinase 2 or map kinase kinase two or map2k2 or mek2 or mek 2 or erk2 or erk 2).mp,kw.	6331

7	(mapk adj5 (kinase 2 or kinase two)).mp,kw.	373

8	Proto-Oncogene Proteins A-raf/	82

9	(araf or raf1 or raf 1).mp,kw.	5081

10	exp ras Proteins/	23921

11	(nras or n ras or kras or k ras or hras or h ras).mp,kw.	33474

12	(ras adj4 (gtpase? or gene* or protein*)).mp,kw.	46270

13	(p21 or cras or c ras).mp,kw.	51017

14	Neurofibromatosis 1/	10525

15	Genes, Neurofibromatosis 1/	928

16	Neurofibromin 1/	1475

17	(neurofibromatos#s 1 or neurofibromatos#s one or recklinghausen* or watson syndrome or neurofibromin).mp,kw.	14177

18	((nf1 or nf 1) adj3 protein).mp,kf.	422

19	Mitogen-Activated Protein Kinase 3/	14310

20	Mitogen-Activated Protein Kinase 1/	14713

21	(mitogen activated protein kinase 1 or mitogen activated protein kinase one or map kinase 1 or map kinase one or mitogen activated protein kinase 3 or mitogen activated protein kinase three or map kinase 3 or map kinase three or mapk1 or mapk 1 or mapk3 or mapk 3).mp,kw.	19169

22	MAP Kinase Signaling System/	36965

23	Mitogen-Activated Protein Kinase Kinases/	8600

24	or/1-23	188338

25	gefitinib/	4903

26	gefitinib.mp,kw.	8146

27	cetuximab/	5157

28	(cetuximab* or erbitux).mp,kw.	8426

29	exp trastuzumab/	8350

30	(trastuzumab or herceptin or trazimera or adotrastuzumab).mp,kw.	14091

31	Vemurafenib/	1558

32	(vemurafenib or zelboraf).mp,kw.	2763

33	Alectinib.mp,kw.	873

34	Cabozantinib.mp,kw.	1399

35	(XL184 or XL 184).mp,kw.	94

36	BMS 907351.mp,kw.	5

37	Crizotinib/	1610

38	(crizotinib or xalkori or 53ah36668s or "pf 02341066" or pf 2341066 or pf-02341066 or pf-2341066 or pf02341066 or pf2341066).mp,kw.	3136

39	Brigatinib.mp,kw.	298

40	(AP26113 or AP 26113).mp,kw.	45

41	Ceritinib.mp,kw.	633

42	(LDK378 or LDK 378).mp,kw.	49

43	Larotrectinib.mp,kw.	285

44	(LOXO 101 or LOXO101).mp,kw.	23

45	RXDX-101.mp,kw.	16

46	(lapatinib or tykerb or 0vua21238f or g873gx646r or gw 282974x or gw 572016 or gw282974x or gw572016).mp,kw.	3137

47	Tucatinib.mp,kw.	111

48	(ONT 380 or ARRY380 or ARRY 380).mp,kw.	10

49	Almonertinib.mp,kw.	29

50	Aumolertinib.mp,kw.	25

51	Sotorasib.mp,kw.	135

52	Adagrasib.mp,kw.	61

53	Pimasertib.mp,kw.	47

54	(AMG510 or AMG 510).mp,kw.	69

55	(MRTX849 or MRTX 849).mp,kw.	44

56	(map* adj3 inhibitor?).mp,kw.	10101

57	(mek* adj3 inhibitor?).mp,kw.	9974

58	(erk* adj3 inhibitor?).mp,kw.	8185

59	(raf adj3 inhibitor?).mp,kw.	1806

60	(braf* adj3 inhibitor?).mp,kw.	3733

61	(b-raf* adj3 inhibitor?).mp,kw.	353

62	mapk?.mp,kw.	78051

63	mek?.mp,kw.	26148

64	erk?.mp,kw.	87115

65	map*kinase?.mp,kw.	1234

66	mapkkk*.mp,kw.	686

67	mapk-erk?.mp,kw.	5038

68	mapk*erk*.mp,kw.	391

69	mekk?.mp,kw.	1245

70	map?k?.mp,kw.	83317

71	pan-raf.mp,kw.	83

72	panraf.mp,kw.	3

73	afatinib*.mp,kw.	1928

74	bibw 2992.mp,kw.	60

75	bibw2992.mp,kw.	26

76	gilotrif*.mp,kw.	14

77	giotrif*.mp,kw.	10

78	41ud74l59m.rn.	941

79	erlotinib*.mp,kw.	7700

80	cp 358774.mp,kw.	4

81	cp358774.mp,kw.	2

82	nsc 718781.mp,kw.	1

83	osi 774.mp,kw.	104

84	osi774.mp,kw.	1

85	r 1415.mp,kw.	1

86	tarceva*.mp,kw.	331

87	da87705x9k.rn.	4298

88	iressa*.mp,kw.	817

89	zd 1839.mp,kw.	43

90	zd1839.mp,kw.	448

91	s65743jhbs.rn.	4903

92	osimertinib*.mp,kw.	2178

93	azd 9291.mp,kw.	11

94	azd9291.mp,kw.	215

95	tagrisso*.mp,kw.	32

96	c225.mp,kw.	502

97	c 225.mp,kw.	81

98	pqx0d8j21j.rn.	5157

99	abp 980.mp,kw.	13

100	abp980.mp,kw.	2

101	aryotrust*.mp,kw.	1

102	"bcd 022".mp,kw.	3

103	ct p6.mp,kw.	18

104	ctp6.mp,kw.	1

105	da 3111.mp,kw.	1

106	dmb 3111.mp,kw.	1

107	hd 201.mp,kw.	3

108	hd201.mp,kw.	4

109	hertraz*.mp,kw.	1

110	herzuma*.mp,kw.	12

111	hlx02.mp,kw.	5

112	kanjinti*.mp,kw.	8

113	myl 1401o.mp,kw.	8

114	ogivri*.mp,kw.	8

115	ontruzant*.mp,kw.	10

116	"pf 05280014".mp,kw.	13

117	r 597.mp,kw.	16

118	r597.mp,kw.	5

119	sb 3.mp,kw.	212

120	sb3.mp,kw.	351

121	trasturel*.mp,kw.	2

122	"tx 05".mp,kw.	5

123	tx05.mp,kw.	1

124	vivitra*.mp,kw.	1

125	zedora*.mp,kw.	4

126	zrc 3256.mp,kw.	1

127	p188anx8ck.rn.	8249

128	belvarafenib*.mp,kw.	4

129	lxh254.mp,kw.	6

130	binimetinib*.mp,kw.	293

131	mek 162.mp,kw.	11

132	mek162.mp,kw.	67

133	mektovi*.mp,kw.	6

134	cobimetinib*.mp,kw.	409

135	cotellic*.mp,kw.	5

136	"gdc 0973".mp,kw.	25

137	gdc-0973.mp,kw.	25

138	rg 7420.mp,kw.	2

139	xl 518.mp,kw.	1

140	xl518.mp,kw.	5

141	dabrafenib*.mp,kw.	1580

142	gsk 2118436?.mp,kw.	3

143	gsk2118436?.mp,kw.	33

144	tafinlar*.mp,kw.	16

145	encorafenib*.mp,kw.	254

146	braftovi*.mp,kw.	10

147	lgx 818.mp,kw.	3

148	lgx818.mp,kw.	20

149	trametinib*.mp,kw.	1877

150	gsk 1120212?.mp,kw.	6

151	gsk1120212?.mp,kw.	56

152	jtp 74057.mp,kw.	6

153	mekinist*.mp,kw.	12

154	ulixertinib*.mp,kw.	36

155	bvd 523.mp,kw.	10

156	plx 4032.mp,kw.	26

157	plx4032.mp,kw.	194

158	rg 7204.mp,kw.	1

159	rg7204.mp,kw.	22

160	ro 5185426.mp,kw.	1

161	ro5185426.mp,kw.	2

162	207smy3fqt.rn.	1558

163	or/25-162	197203

164	24 and 163	81782

165	164 not (exp animals/ not humans.sh.)	54292

166	limit 165 to yr="2010 -Current"	38759

[Embase <1996 to 2022 September 15>](https://proxy.library.mcgill.ca/login?url=http://ovidsp.ovid.com/ovidweb.cgi?T=JS&NEWS=N&PAGE=main&SHAREDSEARCHID=5upra9y75n1naRklnVFMTGuNzAYpx5QWFPuTsvTWgFxZ3Vj2Dkar7gxGPvARFhbwF)

1	Raf protein/	10273

2	(craf or c raf).mp,kw.	2231

3	mitogen activated protein kinase kinase 1/	5130

4	(mitogen activated protein kinase kinase 1 or mitogen activated protein kinase kinase one or map kinase kinase 1 or map kinase kinase one or map2k1 or mek1 or mek 1 or mkk1 or mkk 1 or erk1 or erk 1 or "mek1/2").mp,kw.	57866

5	mitogen activated protein kinase kinase 2/	2850

6	(mitogen activated protein kinase kinase 2 or mitogen activated protein kinase kinase two or kinase activator kinase or mek2 or map kinase kinase 2 or map kinase kinase two or map2k2 or mek2 or mek 2 or erk2 or erk 2).mp,kw.	9080

7	(mapk adj5 (kinase 2 or kinase two)).mp,kw.	430

8	A Raf kinase/	198

9	(araf or raf1 or raf 1).mp,kw.	5735

10	Ras protein/	25284

11	oncogene n ras/	8403

12	K ras protein/	26361

13	oncogene h ras/	3452

14	protein p21/	23544

15	(nras or n ras or kras or k ras or hras or h ras).mp,kw.	66935

16	(ras adj4 (gtpase? or gene* or protein*)).mp,kw.	65244

17	(p21 or cras or c ras).mp,kw.	53098

18	exp neurofibromatosis type 1/	4922

19	neurofibromin/	2870

20	(neurofibromatos#s 1 or neurofibromatos#s one or recklinghausen* or watson syndrome or neurofibromin).mp,kw.	6231

21	((nf1 or nf 1) adj3 protein).mp,kf.	470

22	Mitogen-Activated Protein Kinase 3/	44950

23	mitogen activated protein kinase 1/	59950

24	(mitogen activated protein kinase 1 or mitogen activated protein kinase one or map kinase 1 or map kinase one or mitogen activated protein kinase 3 or mitogen activated protein kinase three or map kinase 3 or map kinase three or mapk1 or mapk 1 or mapk3 or mapk 3).mp,kw.	66021

25	mapk signaling/	22176

26	mitogen activated protein kinase kinase/	6098

27	or/1-26	262192

28	gefitinib/	27518

29	gefitinib.mp,kw.	28518

30	exp cetuximab/	31865

31	(cetuximab* or erbitux).mp,kw.	33012

32	exp trastuzumab/	45937

33	(trastuzumab or herceptin or trazimera or adotrastuzumab).mp,kw.	48244

34	vemurafenib/	9071

35	(vemurafenib or zelboraf).mp,kw.	9493

36	alectinib/	2709

37	Alectinib.mp,kw.	2810

38	cabozantinib/	5667

39	Cabozantinib.mp,kw.	5911

40	(XL184 or XL 184).mp,kw.	785

41	BMS 907351.mp,kw.	59

42	Crizotinib/	9999

43	(crizotinib or xalkori or 53ah36668s or "pf 02341066" or pf 2341066 or pf-02341066 or pf-2341066 or pf02341066 or pf2341066).mp,kw.	10666

44	brigatinib/	1251

45	Brigatinib.mp,kw.	1287

46	(AP26113 or AP 26113).mp,kw.	279

47	ceritinib/	2396

48	Ceritinib.mp,kw.	2509

49	(LDK378 or LDK 378).mp,kw.	289

50	larotrectinib/	916

51	Larotrectinib.mp,kw.	1005

52	(LOXO 101 or LOXO101).mp,kw.	107

53	RXDX-101.mp,kw.	103

54	(lapatinib or tykerb or 0vua21238f or g873gx646r or gw 282974x or gw 572016 or gw282974x or gw572016).mp,kw.	14052

55	tucatinib/	538

56	Tucatinib.mp,kw.	552

57	(ONT 380 or ARRY380 or ARRY 380).mp,kw.	98

58	Almonertinib.mp,kw.	58

59	aumolertinib/	52

60	aumolertinib.mp,kw.	52

61	sotorasib/	393

62	sotorasib.mp,kw.	422

63	adagrasib/	187

64	adagrasib.mp,kw.	196

65	pimasertib/	313

66	pimasertib.mp,kw.	319

67	(AMG510 or AMG 510).mp,kw.	250

68	(MRTX849 or MRTX 849).mp,kw.	159

69	(map* adj3 inhibitor?).mp,kw.	12635

70	(mek* adj3 inhibitor?).mp,kw.	15350

71	(erk* adj3 inhibitor?).mp,kw.	10576

72	(raf adj3 inhibitor?).mp,kw.	5265

73	(braf* adj3 inhibitor?).mp,kw.	7200

74	(b-raf* adj3 inhibitor?).mp,kw.	3045

75	mapk?.mp,kw.	109578

76	mek?.mp,kw.	36516

77	erk?.mp,kw.	114839

78	map*kinase?.mp,kw.	4435

79	mapkkk*.mp,kw.	704

80	mapk-erk?.mp,kw.	6515

81	mapk*erk*.mp,kw.	624

82	mekk?.mp,kw.	1342

83	map?k?.mp,kw.	113867

84	pan-raf.mp,kw.	184

85	panraf.mp,kw.	14

86	afatinib/	7228

87	afatinib*.mp,kw.	7503

88	bibw 2992.mp,kw.	594

89	bibw2992.mp,kw.	75

90	gilotrif*.mp,kw.	183

91	giotrif*.mp,kw.	78

92	41ud74l59m.rn.	0

93	erlotinib/	30360

94	erlotinib*.mp,kw.	31408

95	cp 358774.mp,kw.	117

96	cp358774.mp,kw.	2

97	nsc 718781.mp,kw.	6

98	osi 774.mp,kw.	1085

99	osi774.mp,kw.	13

100	r 1415.mp,kw.	6

101	tarceva*.mp,kw.	4035

102	da87705x9k.rn.	0

103	iressa*.mp,kw.	5147

104	zd 1839.mp,kw.	2017

105	zd1839.mp,kw.	565

106	s65743jhbs.rn.	0

107	osimertinib/	5998

108	osimertinib*.mp,kw.	6270

109	azd 9291.mp,kw.	460

110	azd9291.mp,kw.	421

111	tagrisso*.mp,kw.	206

112	c225.mp,kw.	916

113	c 225.mp,kw.	616

114	pqx0d8j21j.rn.	0

115	abp 980.mp,kw.	56

116	abp980.mp,kw.	4

117	aryotrust*.mp,kw.	3

118	"bcd 022".mp,kw.	22

119	ct p6.mp,kw.	50

120	ctp6.mp,kw.	4

121	da 3111.mp,kw.	1

122	dmb 3111.mp,kw.	4

123	hd 201.mp,kw.	6

124	hd201.mp,kw.	9

125	hertraz*.mp,kw.	7

126	herzuma*.mp,kw.	61

127	hlx02.mp,kw.	13

128	kanjinti*.mp,kw.	45

129	myl 1401o.mp,kw.	32

130	ogivri*.mp,kw.	54

131	ontruzant*.mp,kw.	46

132	"pf 05280014".mp,kw.	56

133	r 597.mp,kw.	28

134	r597.mp,kw.	10

135	sb 3.mp,kw.	233

136	sb3.mp,kw.	454

137	trasturel*.mp,kw.	4

138	"tx 05".mp,kw.	11

139	tx05.mp,kw.	5

140	vivitra*.mp,kw.	5

141	zedora*.mp,kw.	12

142	zrc 3256.mp,kw.	1

143	p188anx8ck.rn.	0

144	belvarafenib/	25

145	belvarafenib*.mp,kw.	26

146	lxh254.mp,kw.	15

147	binimetinib/	1541

148	binimetinib*.mp,kw.	1585

149	mek 162.mp,kw.	261

150	mek162.mp,kw.	173

151	mektovi*.mp,kw.	40

152	cobimetinib/	2108

153	cobimetinib*.mp,kw.	2175

154	cotellic*.mp,kw.	66

155	"gdc 0973".mp,kw.	226

156	gdc-0973.mp,kw.	226

157	rg 7420.mp,kw.	5

158	xl 518.mp,kw.	63

159	xl518.mp,kw.	7

160	dabrafenib/	6042

161	dabrafenib*.mp,kw.	6387

162	gsk 2118436?.mp,kw.	295

163	gsk2118436?.mp,kw.	112

164	tafinlar*.mp,kw.	245

165	encorafenib/	1137

166	encorafenib*.mp,kw.	1201

167	braftovi*.mp,kw.	46

168	lgx 818.mp,kw.	139

169	lgx818.mp,kw.	63

170	trametinib/	7656

171	trametinib*.mp,kw.	7884

172	gsk 1120212?.mp,kw.	394

173	gsk1120212?.mp,kw.	192

174	jtp 74057.mp,kw.	28

175	mekinist*.mp,kw.	254

176	ulixertinib*.mp,kw.	196

177	bvd 523.mp,kw.	71

178	plx 4032.mp,kw.	914

179	plx4032.mp,kw.	480

180	rg 7204.mp,kw.	91

181	rg7204.mp,kw.	60

182	ro 5185426.mp,kw.	51

183	ro5185426.mp,kw.	13

184	207smy3fqt.rn.	0

185	or/28-184	342381

186	27 and 185	120825

187	186 not ((exp animal/ or animal experiment/ or nonhuman/) not (exp human/ or human experiment/))	80965

188	limit 187 to yr="2010 -Current"	66686
