## Supplementary material for "Clinical activity of Mitogen-Activated Protein Kinase (MAPK) inhibitors in patients with MAP2K1 (MEK1)-mutated metastatic cancers": Figure S1

### Slide 1
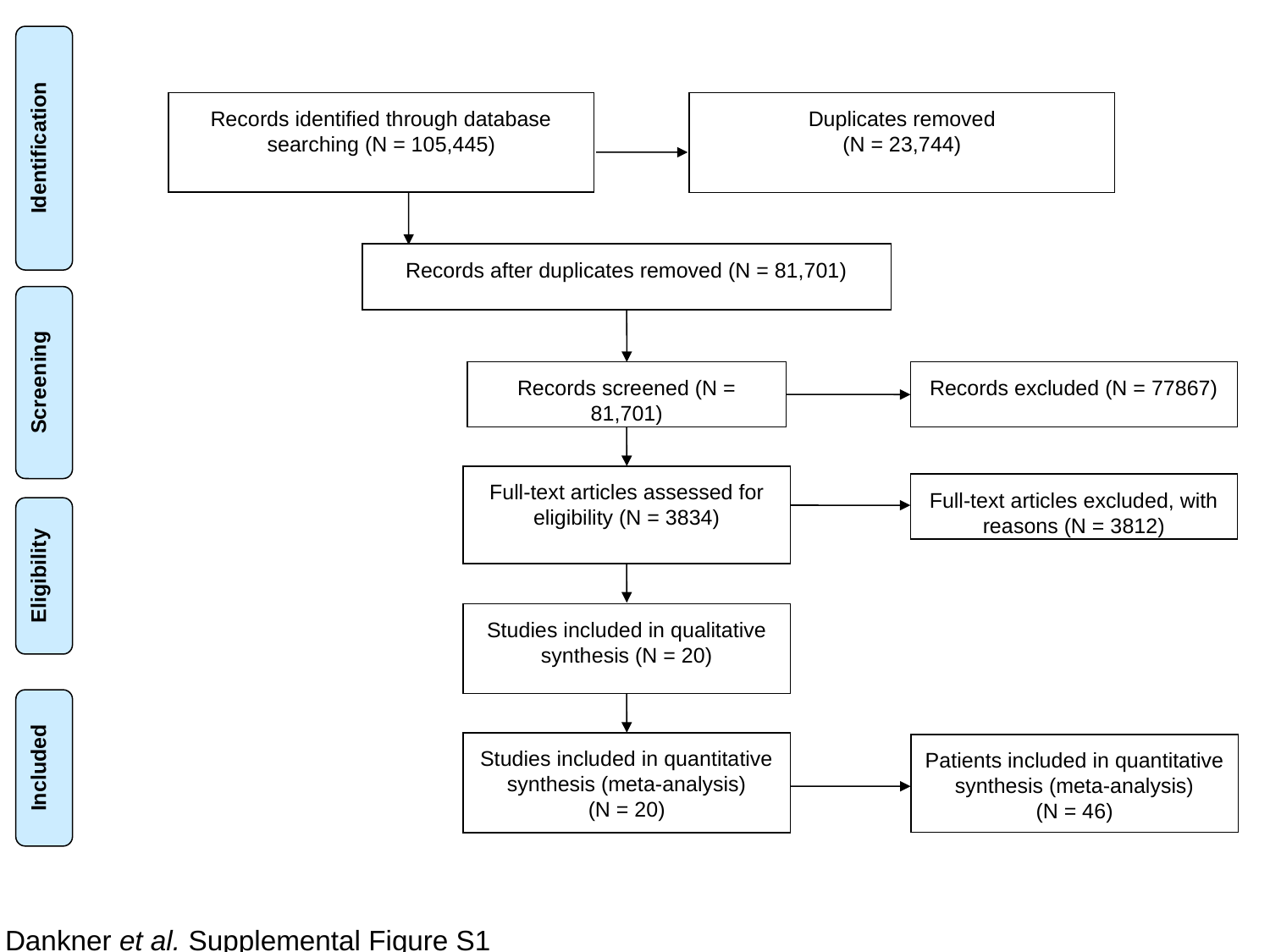

Records identified through database searching (N = 105,445)
Duplicates removed
(N = 23,744)
Identification
Records after duplicates removed (N = 81,701)
Screening
Records screened (N = 81,701)
Records excluded (N = 77867)
Full-text articles assessed for eligibility (N = 3834)
Full-text articles excluded, with reasons (N = 3812)
Eligibility
Studies included in qualitative synthesis (N = 20)
Included
Studies included in quantitative synthesis (meta-analysis)
(N = 20)
Patients included in quantitative synthesis (meta-analysis)
(N = 46)
Dankner et al. Supplemental Figure S1
