## Supplementary material for "Clinical activity of Mitogen-Activated Protein Kinase (MAPK) inhibitors in patients with MAP2K1 (MEK1)-mutated metastatic cancers": Figure S2

### Slide 1
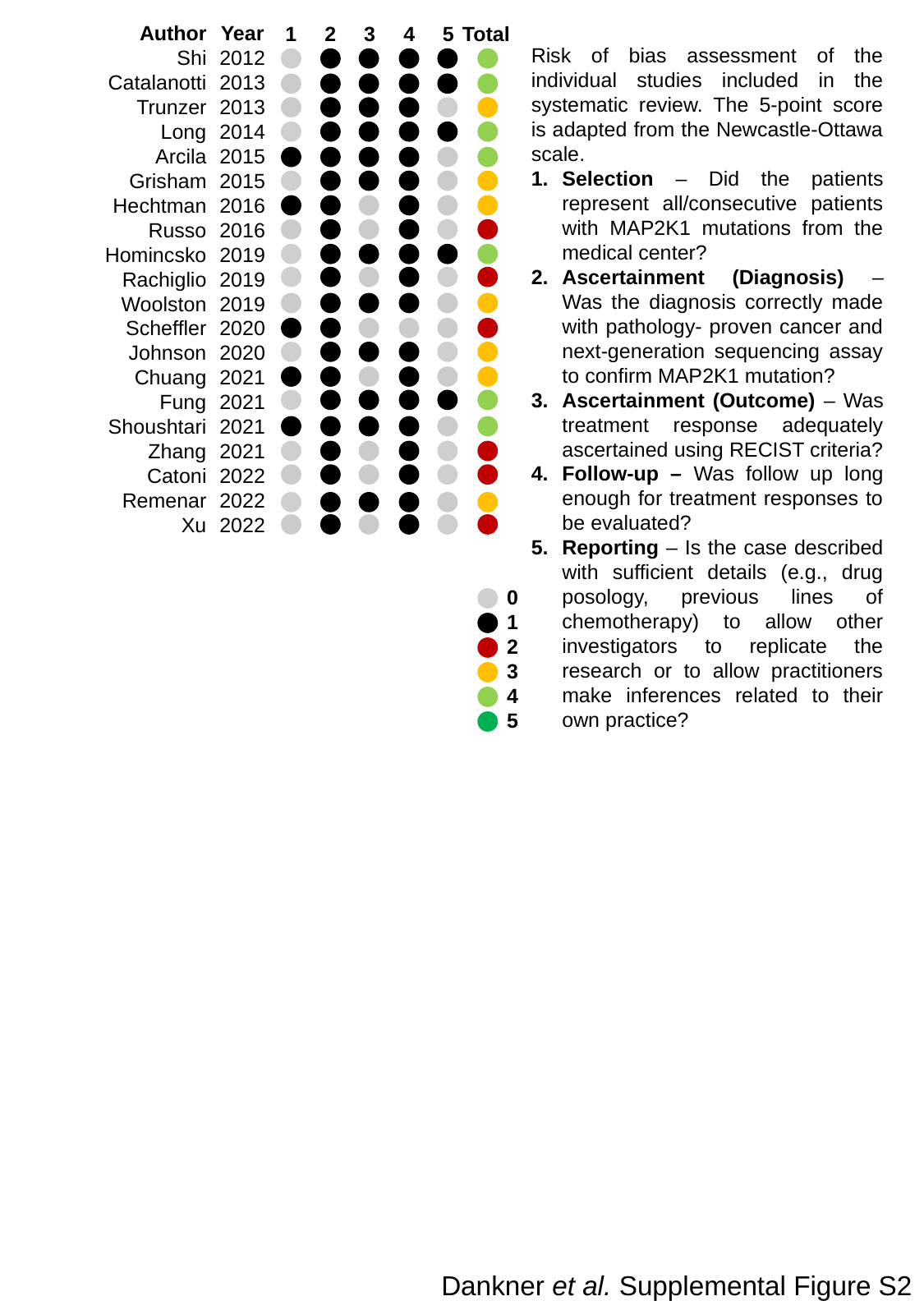

Author
Shi
Catalanotti
Trunzer
Long
Arcila
Grisham
Hechtman
Russo
Homincsko
Rachiglio
Woolston
Scheffler
Johnson
Chuang
Fung
Shoushtari
Zhang
Catoni
Remenar
Xu
Year
2012
2013
2013
2014
2015
2015
2016
2016
2019
2019
2019
2020
2020
2021
2021
2021
2021
2022
2022
2022
Total
4
5
1
3
2
Risk of bias assessment of the individual studies included in the systematic review. The 5-point score is adapted from the Newcastle-Ottawa scale.
Selection – Did the patients represent all/consecutive patients with MAP2K1 mutations from the medical center?
Ascertainment (Diagnosis) – Was the diagnosis correctly made with pathology- proven cancer and next-generation sequencing assay to confirm MAP2K1 mutation?
Ascertainment (Outcome) – Was treatment response adequately ascertained using RECIST criteria?
Follow-up – Was follow up long enough for treatment responses to be evaluated?
Reporting – Is the case described with sufficient details (e.g., drug posology, previous lines of chemotherapy) to allow other investigators to replicate the research or to allow practitioners make inferences related to their own practice?
0
1
2
3
4
5
Dankner et al. Supplemental Figure S2
