## Supplementary material for "Clinical activity of Mitogen-Activated Protein Kinase (MAPK) inhibitors in patients with MAP2K1 (MEK1)-mutated metastatic cancers": Figure S3

### Slide 1
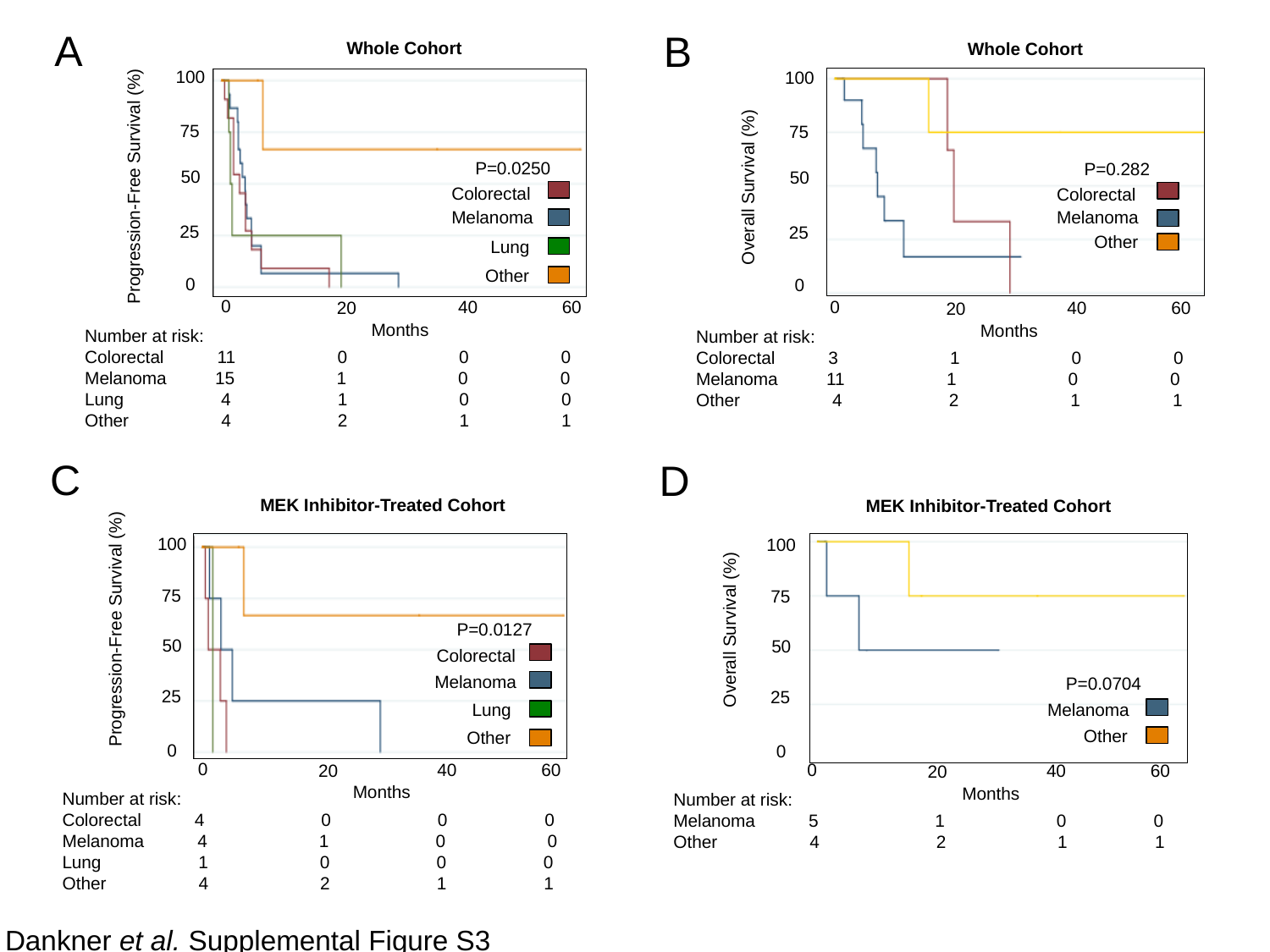

A
Whole Cohort
100
75
P=0.0250
50
Progression-Free Survival (%)
Colorectal
Melanoma
25
Lung
Other
0
0
60
40
20
Months
Number at risk:
Colorectal 11 0 0 0
Melanoma 15 1 0 0
Lung 4 1 0 0
Other 4 2 1 1
C
MEK Inhibitor-Treated Cohort
100
75
P=0.0127
Progression-Free Survival (%)
50
Colorectal
Melanoma
25
Lung
Other
0
0
60
40
20
Months
Number at risk:
Colorectal 4 0 0 0
Melanoma 4 1 0 0
Lung 1 0 0 0
Other 4 2 1 1
B
Whole Cohort
100
75
P=0.282
50
Overall Survival (%)
Colorectal
Melanoma
25
Other
0
0
60
40
20
Months
Number at risk:
Colorectal 3 1 0 0
Melanoma 11 1 0 0
Other 4 2 1 1
D
MEK Inhibitor-Treated Cohort
100
75
Overall Survival (%)
50
P=0.0704
25
Melanoma
Other
0
0
60
40
20
Months
Number at risk:
Melanoma 5 1 0 0
Other 4 2 1 1
Dankner et al. Supplemental Figure S3
