## Supplementary material for "Clinical activity of Mitogen-Activated Protein Kinase (MAPK) inhibitors in patients with MAP2K1 (MEK1)-mutated metastatic cancers": Figure S4

### Slide 1
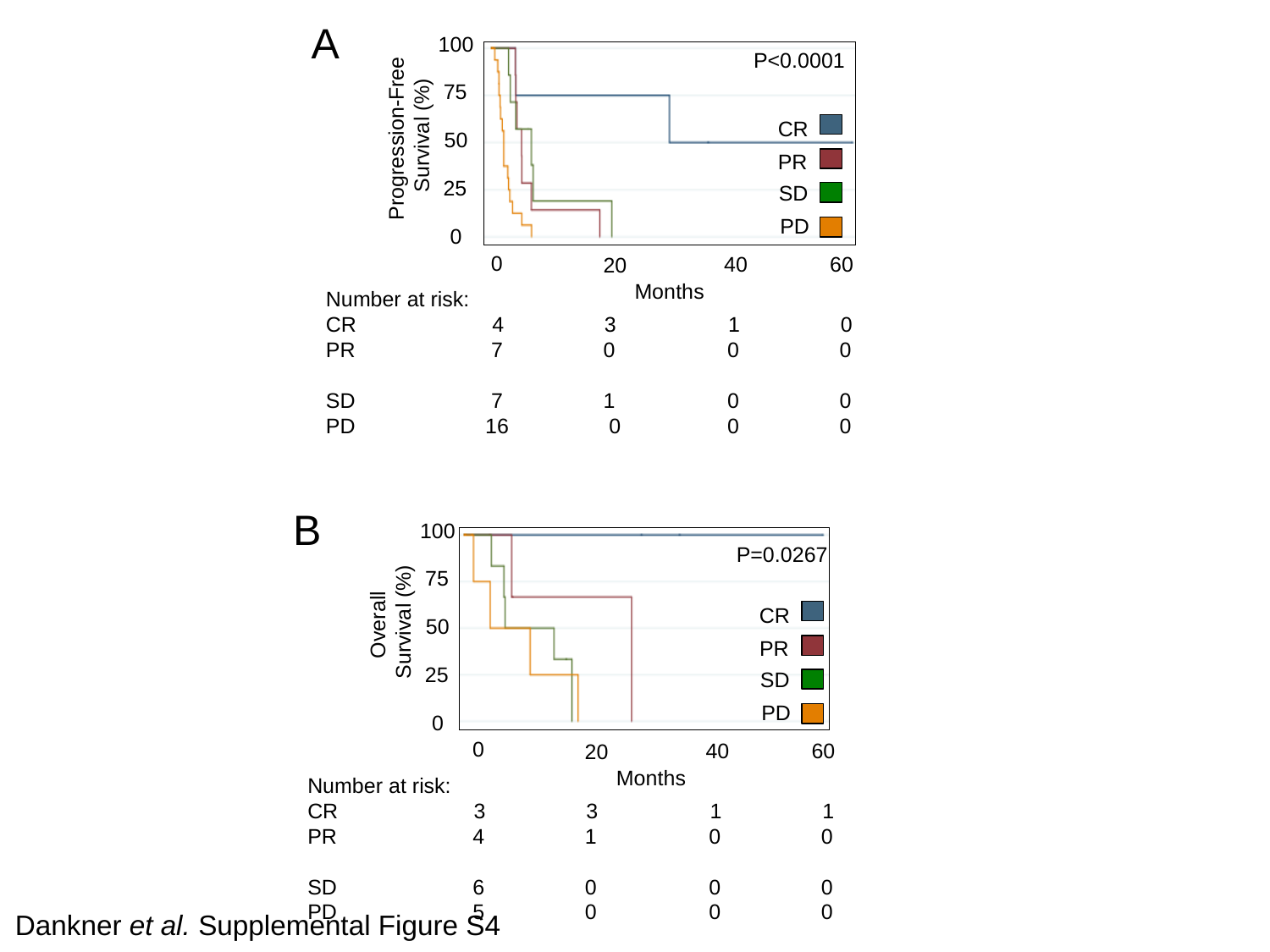

A
100
P<0.0001
75
Progression-Free
Survival (%)
CR
50
PR
25
SD
PD
0
0
60
40
20
Months
Number at risk:
CR 4 3 1 0
PR 7 0 0 0
SD 7 1 0 0
PD 16 0 0 0
B
100
P=0.0267
75
Overall
Survival (%)
CR
50
PR
25
SD
PD
0
0
60
40
20
Months
Number at risk:
CR 3 3 1 1
PR 4 1 0 0
SD 6 0 0 0
PD 5 0 0 0
Dankner et al. Supplemental Figure S4
