## Supplementary material for "Clinical activity of Mitogen-Activated Protein Kinase (MAPK) inhibitors in patients with MAP2K1 (MEK1)-mutated metastatic cancers": Figure S5

### Slide 1
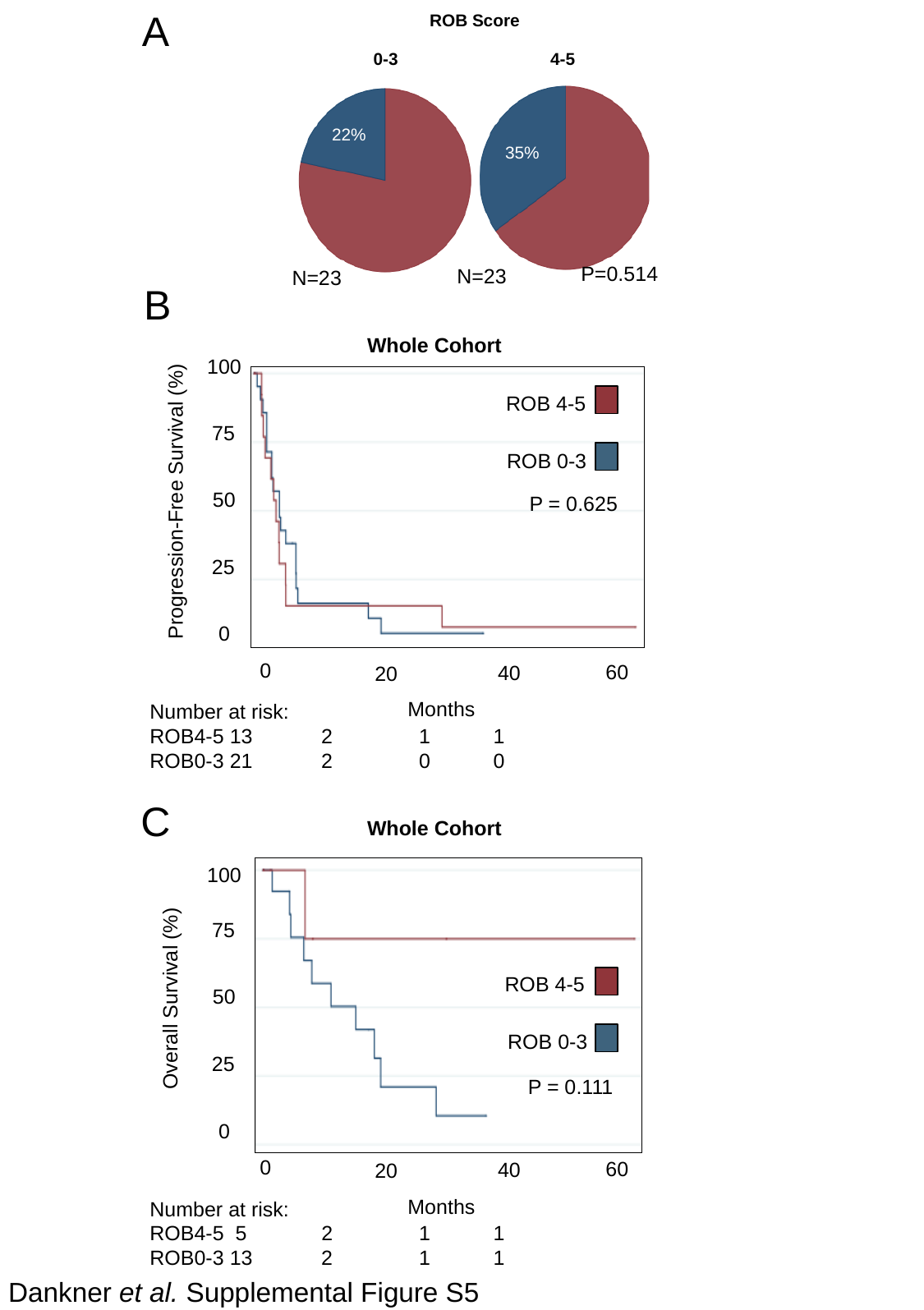

A
ROB Score
4-5
0-3
22%
35%
P=0.514
N=23
N=23
B
Whole Cohort
100
ROB 4-5
75
ROB 0-3
Progression-Free Survival (%)
50
25
0
0
60
40
20
Months
Number at risk:
ROB4-5 13 2 1 1
ROB0-3 21 2 0 0
P = 0.625
C
Whole Cohort
100
75
ROB 4-5
Overall Survival (%)
50
ROB 0-3
25
P = 0.111
0
0
60
40
20
Months
Number at risk:
ROB4-5 5 2 1 1
ROB0-3 13 2 1 1
Dankner et al. Supplemental Figure S5
