## Supplementary material for "Clinical activity of Mitogen-Activated Protein Kinase (MAPK) inhibitors in patients with MAP2K1 (MEK1)-mutated metastatic cancers": Table S1

| Treatment Category | Drug Names | Number of Patients |
|---|---|---|
| MEKi | Trametinib | 7 |
| | Binimetinib | 7 |
| | Selumetinib | 4 |
| | FDA-Approved MEKi, unspecified | 1 |
| MEKi + BRAFi | Vemurafenib + Cobimetinib | 1 |
| | Dabrafenib + Trametinib | 1 |
| MEKi + EGFRi | Panitimumab + Trametinib | 1 |
| MEKi + EGFRi + BRAFi | Cetuximab + Encorafenib + Binimetinib | 2 |
| | Cetuximab + Dabrafenib + Trametinib | 1 |
| BRAFi | Vemurafenib | 8 |
| | Dabrafenib | 3 |
| EGFRi | Erlotinib | 3 |
| | Panitimumab | 4 |
| | Cetuximab | 3 |

**Supplemental Table S1:** MAPK targeted therapy regimens used for patients in the meta-analysis.
