## Supplementary material for "Clinical activity of Mitogen-Activated Protein Kinase (MAPK) inhibitors in patients with MAP2K1 (MEK1)-mutated metastatic cancers": Table S2

| | # of Patients | # of Patients with Response | Response Rate | Odds Ratio | 95% Confidence Interval | P-Value |
|---|---|---|---|---|---|---|
| *Entire Cohort* | 46 | 13 | 28.3% | | | |
| *Study Characteristics* | | | | | | |
| **Study Type** | | | | | | |
| Prospective | 21 | 4 | 19.0% | 0.418 | 0.107 - 1.632 | 0.21 |
| Retrospective | 25 | 9 | 36.0% | | | |
| **Response Criteria** | | | | | | |
| RECIST | 32 | 11 | 34.4% | 3.143 | 0.594 - 16.616 | 0.178 |
| non-RECIST | 14 | 2 | 14.3% | | | |
| **Geographic Location** | | | | | | |
| North America | 37 | 11 | 29.7% | 1.481 | 0.265 -8.288 | 0.655 |
| Other | 9 | 2 | 22.2% | | | |
| *Patient & Treatment Characteristics* | | | | | | |
| **Gender** | | | | | | |
| Male | 21 | 6 | 28.6% | 0.933 | 0.243 - 3.582 | 0.92 |
| Female | 20 | 6 | 30.0% | | | |
| **Age** | | | | | | |
| >/= 65 | 12 | 2 | 16.7% | 0.400 | 0.0719 - 2.225 | 0.295 |
| < 65 | 27 | 9 | 33.3% | | | |
| **Cancer Type** | | | | | | |
| Melanoma | 19 | 7 | 36.8% | 2.041 | 0.556 - 7.497 | 0.282 |
| Colrectal Cancer | 12 | 2 | 16.7% | 0.418 | 0.078 - 2.244 | 0.309 |
| Lung Cancer | 11 | 2 | 18.2% | 0.485 | 0.0894 - 2.628 | 0.401 |
| Other | 4 | 2 | 50.0% | 2.817 | 0.353 - 22.487 | 0.328 |
| **Therapy Type** | | | | | | |
| MEKi-containing regimen | 24 | 7 | 29.2% | 1.098 | 0.303 - 3.975 | 0.887 |
| Non-MEKi-containing regimen | 22 | 6 | 27.2% | | | |
| **Concomitant Chemotherapy** | | | | | | |
| Yes | 13 | 3 | 23.1% | 0.690 | 0.156 - 3.057 | 0.625 |
| No | 33 | 10 | 30.3% | | | |
| **Prior Lines of Therapy** | | | | | | |
| 0 | 11 | 4 | 36.4% | 0.636 | 0.119 - 3.411 | 0.598 |
| 1+ | 15 | 4 | 26.7% | | | |
| **Co-Occuring MAPK Mutation** | | | | | | |
| Present | 25 | 7 | 28.0% | 0.972 | 0.268 - 3.525 | 0.966 |
| Absent | 21 | 6 | 28.6% | | | |
| **Co-Occuring BRAF V600 Mutation** | | | | | | |
| Present | 18 | 6 | 33.0% | 2.733 | 0.343 - 21.763 | 0.342 |
| Absent | 28 | 7 | 25.0% | | | |
| **Co-Occuring non-MAPK Oncogenic Mutation** | | | | | | |
| Present | 17 | 4 | 23.5% | 0.684 | 0.174 - 2.689 | 0.586 |
| Absent | 29 | 9 | 31.0% | | | |
| **MAP2K1 Class** | | | | | | |
| Unclassified | 3 | 1 | 33.3% | 1.292 | 0.107 - 15.604 | 0.840 |
| 1 | 15 | 4 | 26.7% | 0.890 | 0.223 - 3.543 | 0.868 |
| 2 | 26 | 8 | 30.8% | 1.333 | 0.359 - 4.945 | 0.667 |
| 3 | 2 | 0 | 0.0% | <0.001 | NA | **0.009** |

**Supplemental Table S2: Overall response rates associated with clinical variables**
